# Sugar or Salt (“SOS”): a protocol for a UK multicentre randomised trial of mannitol and hypertonic saline in severe traumatic brain injury and intracranial hypertension

**DOI:** 10.1101/19008276

**Authors:** MJ Rowland, T Veenith, C Scomparin, MH Wilson, PJ Hutchinson, AG Kolias, R Lall, S Regan, J Mason, PJD Andrews, D Horner, J Naisbitt, A Devrell, A Malins, P Dark, DF McAuley, GD Perkins

## Abstract

Hyperosmolar solutions are widely used to treat raised intracranial pressure (ICP) following severe traumatic brain injury (TBI). Although mannitol has historically been the most frequently administered, hypertonic saline (HTS) solutions are increasingly being used. However, definitive evidence regarding their comparative effectiveness is lacking. The Sugar or Salt (SOS) Trial is a UK randomised, allocation concealed open label multicentre pragmatic trial designed to determine the clinical and cost-effectiveness of hypertonic saline (HTS) compared with mannitol in the management of patients with severe TBI. Patients requiring intensive care unit admission and intracranial pressure (ICP) monitoring post-TBI will be allocated at random to receive equi-osmolar boluses of either mannitol or HTS following failure of routine first line measures to control ICP. The primary outcome for the study will be the Extended Glasgow Outcome Scale (GOS-E) assessed at 6 months after randomisation. Results will inform current clinical practice in the routine use of hyperosmolar therapy as well as assess the impact of potential side effects. Pre-planned longer term clinical and cost effectiveness analyses will further inform the use of these treatments.

## Background

Traumatic brain injury (TBI) is a major cause of death and severe disability with sixty-nine million individuals worldwide estimated to sustain a TBI each year.^1^ In the United Kingdom, approximately 3,500 patients have moderate to severe TBI requiring treatment in intensive care units (ICUs) each year.^2^ Resource use in these patients is high with an average length of stay in ICU of 9 days.^2^ Long term outcomes from TBI remain poor – with reported mortality as high as 26% at 6 months and significant disability in survivors.^3^

Following TBI, intracranial pressure (ICP) may be increased by a mass effect from intracranial haematomas, contusions, diffuse brain swelling, or hydrocephalus. A number of studies have shown an association between raised intracranial pressure (ICP) and poor neurological outcomes.^4,5^ As a result, the treatment of elevated ICP has been a central focus of both the medical and surgical management of patients with severe TBI on the ICU.

The intensive care management of patients with severe TBI and raised ICP usually takes a stepwise approach. Patients receive “stage 1” interventions (sedation, artificial ventilation, blood pressure and temperature control, and head-up positioning) as part of routine ICU care. If these measures fail to limit ICP to approximately 20-25mmHg, medical management with either the drug mannitol (a sugar alcohol) or hypertonic saline (HTS) is used alongside other “stage 2” measures. If these drugs (osmotherapy) fail to control ICP, “stage 3” measures include surgical decompression and barbiturate-induced coma.

Both mannitol and HTS are thought to have a similar core mechanism of action. They both increase the osmotic pressure of plasma in a dose-dependent fashion. The increased plasma osmotic pressure draws water from extracellular spaces in brain tissue, decreasing the volume of both oedematous and normal brain tissue and thereby decreasing ICP. The increased plasma osmolality also draws water into the circulation from all tissue extracellular spaces and so increases circulating volume, cardiac output and possibly blood pressure, whilst decreasing blood viscosity by haemodilution. These effects all increase cerebral blood flow and oxygen delivery. Mannitol also causes an osmotic diuresis with increased free water clearance and a further secondary increase in plasma osmolarity.

While mannitol has been the traditional hyperosmolar agent of choice, use of HTS is increasing. This has been supported by several recent small but heterogeneous, studies.^6-8^ A number of systematic reviews have also been published recently reviewing trials investigating the use of osmotherapy in TBI.^9-11^ The included trials were limited by moderate to high risk of bias, inconsistency, imprecision and indirectness, and spanned three decades during which management of traumatic brain injury evolved. Although reduction in ICP has been consistently demonstrated with both mannitol and HTS, there is a suggestion that HTS provides a more robust and durable effect in lowering ICP.^6,7^

To provide a definitive answer to whether mannitol or HTS is the better agent for osmotherapy, the National Institute for Health Research (NIHR) has commissioned the “Sugar or Salt” (SOS) trial (HTA reference 17/120/01). This is a UK randomised, multi-centre, allocation concealed, open label trial designed to assess the relative effects of mannitol or HTS on long-term neurological outcome when used to treat raised intracranial pressure in patients with traumatic brain injury (TBI). This paper provides a summary of the trial protocol. A full copy of the trial protocol and any subsequent updates can be found at: https://warwick.ac.uk/fac/sci/med/research/ctu/

### Study objectives

#### Primary objective

The primary objective of the SOS Trial is to determine whether hyperosmolar therapy regimes involving hypertonic saline or mannitol improve neurological function at 6 months (as assessed by the GOS-E questionnaire) following severe TBI with raised ICP.

#### Secondary objectives

Secondary objectives are to assess the effects of hyperosmolar therapy with either hypertonic saline or mannitol on clinical, patient-centred and economic outcomes in the ICU, in hospital and up to 12 months follow-up post randomisation. These will address all the outcomes identified in the HTA commissioning brief and provide a definitive assessment of the clinical and cost effectiveness of hyperosmolar treatments.

## Methods and design

This protocol manuscript was written in concordance with the SPIRIT guidelines.^12^

### Trial design

The SOS trial is an allocation concealed, open label, randomised controlled clinical and cost effectiveness trial with an internal pilot and blinded assessment of primary outcome at 6 months. The main trial will be preceded by a six-month internal pilot study in 8-10 ICUs to test recruitment feasibility and compliance with assigned treatment. The progression of the pilot will be informed by the recently published best practice guidelines.^13^ The trial will be conducted and managed by the Warwick Clinical Trials Unit (WCTU) and sponsored by the University Hospitals Birmingham NHS Foundation Trust.

The funding is provided by the National Institute for Health Research (NIHR) following a commissioned call from the Health Technology Assessment programme (HTA study reference 17/120/01). The trial will be conducted in accordance with the principles of the Declaration of Helsinki and Good Clinical Practice.

### Participants, interventions and outcomes

#### Study setting

The main trial will take place in at least 25 UK NHS hospitals with ICUs that routinely manage patients with severe TBI and where medical staff managing the patient have clinical equipoise for the use of protocolised hyperosmolar therapy and agree to maintain trial allocation in randomised patients.

Staff should also demonstrate and document a willingness to comply with the protocol, the principles of GCP (Good Clinical Practice) and regulatory requirements and be prepared to participate in training. Sites will need to establish experience with receiving and acting on protocolised advise in relation to the administration of hyperosmolar therapy. This will be addressed with a ‘run-in’ period while the pilot phase is ongoing with all sites having access to an educational package.

### Eligibility criteria

#### Inclusion criteria

Patients will be included who:

- Are aged ≥16 years old
- Have been admitted to ICU following TBI within 10 days from initial primary head injury
- Have an abnormal CT scan consistent with TBI
- Have an ICP >20mmHg for more than 5 mins despite stage 1 procedures

#### Exclusion criteria

Patients will be excluded if they meet one or more of the following criteria:

- Devastating brain injury with withdrawal of life sustaining treatment anticipated in the next 24 hours
- Pregnancy
- Severe hypernatraemia (serum Na > 155 mmol/L)

The inclusion and exclusion criteria are designed to include those who reflect the general population of patients with severe TBI and raised ICP who may benefit from the therapeutic intervention and exclude those patients who are unlikely to benefit owing to their underlying condition or disease trajectory.

#### Trial interventions

Providing the patient meets the inclusion criteria above, they will be randomly assigned to boluses of either:

- 2ml/kg 20% mannitol intravenous bolus (osmolarity = 1100 mOsm/L) – or equivalent osmolar dose using the concentration used locally by participating study centres
- 2ml/kg 3% hypertonic saline intravenous bolus (osmolarity = 1026 mOsm/L) – or equivalent osmolar dose using the concentration used locally by participating study centres

The IMP will be administered by clinical staff in accordance with local policy. If the ICP remains >20mmHg, boluses of each IMP can be repeated until serum sodium reaches 155 mmol/L. If there is a second spike in ICP to >20mmHg, allocated IMP should continue to be used.

### Outcome measures

#### Primary outcome measure

Extended Glasgow Outcome Scale (GOS-E) measured at 6 months after randomisation

#### Secondary outcome measures

##### Efficacy

1. ICP control (during period of monitoring in ICU)
2. Progression to stage 3 therapies
3. Requirement for stage 3 therapies to control ICP

##### Resource use

4. Organ support requirements during ICU
5. Critical care length of stay
6. Hospital length of stay

##### Patient outcomes

7. Longer term neurological outcomes: Modified Oxford Handicap Scale (mOHS) at discharge and GOS-E at 12 months
8. Survival: to hospital discharge (the time at which the patient is discharged from the hospital regardless of neurological status, outcome or destination) and at 3 months, 6 months and 12 months
9. Quality of life: EQ-5D-5L at hospital discharge, 3 months, 6 months and 12 months post-TBI

### Sample size

The planned size of this trial is 638 patients. This is based on a superiority hypothesis of a difference between the two interventions, using 90% power, two-sided significance level of 5% with a dropout/withdrawal rate of 6%. In order to detect a treatment reduction of 13% (63.5% to 50.5%) in the proportion of patients having an unfavourable neurological outcome (GOS-E: dead, vegetative state, lower severe disability, upper severe disability) compared to favourable outcome (GOS-E: lower moderate disability, upper moderate disability, lower good recovery, upper good recovery) between mannitol and hypertonic saline, 319 patients will be required on each arm.

The sample size was informed by the following evidence. The GOS-E is an 8-point ordinal scale (1= death and 8=upper good recovery). In studies carried out in TBI patients, this scale is often used as a dichotomous outcome, with GOS-E categories 1-4: poor versus 5-8: good.^14-17^ In calculating the sample size for an ordinal scale, there is a requirement for a proportional odds over the categories.^18^ Currently, there is some indication of the violation of this proportional odds assumption (e.g. Hutchinson PJ et al.^16^) and for this reason the sample size using the ordinal approach will have limitations. In light of this, for the purposes of the SOS Trial, the GOS-E has been dichotomised in the conventional way. In addition to this, the binary approach is a more conservative approach and analysis using the ordinal categories will furtherincrease the statistical power of the study.

The proportion of patients with unfavourable neurological outcome on the mannitol arm range from 37%-70% across trials in patients with TBI.^14-17^ For our trial, we have taken the worse outcome as 63.5% which is representative of the larger trial samples as illustrated in the Rescue ICP study (Hutchinson PJ et al., 2016 - 60% in the mannitol arm^16^) and EUROTHERM trial (Andrews PJ et al., 2015 - 63.5% in the mannitol arm^14^).

The clinically important difference ranges from 10% to 20%.^14-17^ Our clinically important reduction of proportion of patients with an unfavourable neurological outcome fits in with an achievable sample size of 638 patients as well as aiming to minimise the difference that would be considered relevant.

Finally, loss of follow-up in UK critical care trials is often low (<3%). The EUROTHERM trial (Andrews PJ et al.^14^) reported a 1% withdrawal rate and the drop-out rate for the Rescue ICP study was 6% (Hutchinson PJ et al.^16^). Many of the studies of patients with TBI reviewed report no withdrawal rates. In line with the Rescue ICP study^16^ we have estimated a drop-out rate of 6% for the SOS Trial.

### Assignment of interventions

#### Randomisation

Patients will be randomised in a ratio of 1:1 to either mannitol or HTS using a secure web-based and allocation concealed randomisation system. In the event that the web-based system cannot be used, an emergency Interactive Voice Response (IVR) randomisation system will also be in place.

#### Blinding

As key clinical parameters (urine output and serum sodium levels) monitored in TBI patients will be influenced by the IMPs, it will not be possible or safe to blind clinical staff on the ICU as to the patient’s treatment allocation. Investigators assessing the primary and secondary endpoints will be blinded to treatment allocation though.

### Data collection, management and analysis

#### Data collection

Data will be entered via an electronic case report form (eCRF). Clinical data will be collected during the ICU stay up to 28 days after randomisation (outlined in the study schematic in **Figure 1** and detailed in **Table 1**). Baseline characteristics collected include patient demographics, comorbidities, pre-admission function, inclusion/exclusion criteria, consent, GCS motor score, date/time and mechanism of TBI, temperature at hospital admission, CT scan appearance (Marshall category 1-6), details of injuries to head and other body systems. Daily data captured following randomisation will include core temperature, ICP, blood pressure, ICU/hospital admission status, resource use (Critical Care Minimum Dataset (CCMDS)), AEs (treatment failure, need for other treatments, electrolytes, renal function, renal replacement therapy), survival status. After discharge, data will be collected on GOS-E, survival status and utilisation of community care resources after acute hospital discharge up to 12 months after randomisation. These data will be collected by telephone questionnaire assessment of either the patient or primary carer. Health related quality of life up to 12 months after randomisation will be collected using the EQ-5D-5L – a generic preference-based measure of health that is recommended by NICE for economic evaluations).

**Table 1:** SOS Trial schedule of interventions.

|  | 1 | 2 | 3 | 4 | 5 | 6 | 7 |
| --- | --- | --- | --- | --- | --- | --- | --- |
|  | ICPM insertion | ICP > 20 for 5 minutes | Hospital | Hospital discharge | Month 3 (2 - 4) | Month 6 (5 - 10) | Month 12 (11-13) |
| Informed consent | X | X | ✓ | X | X | X | X |
| Inclusion/exclusion criteria | ✓ | ✓ | X | X | X | X | X |
| Patient identifiers | ✓ | X | X | X | X | X | X |
| Demographics | ✓ | X | X | X | X | X | X |
| Medical history | ✓ | X | X | X | X | X | X |
| Best GCS score prior to intubation/sedation | ✓ | X | X | X | X | X | X |
| CT scan | ✓ | X | X | X | X | X | X |
| Details of TBI | ✓ | X | X | X | X | X | X |
| Intervention | X | ✓ | X | X | X | X | X |
| ICP control | ✓ | ✓ | ✓ | X | X | X | X |
| Adverse event reporting, stage 3 therapies, treatment failure, organ failure | X | ✓ | ✓ | X | X | X | X |
| ICU/Hospital length of stay | X | ✓ | ✓ | ✓ | X | X | X |
| Survival status | X | X | X | ✓ | ✓ | ✓ | ✓ |
| Neurological outcome (mOHS) | X | X | X | ✓ | X | X | X |
| Neurological outcome (GOS-E) | X | X | X | X | X | ✓ | ✓ |
| Quality of Life (EQ-5D-5L) | X | X | X | ✓ | ✓ | ✓ | ✓ |
| Health Economics Questionnaire | X | X | X | X | ✓ | ✓ | ✓ |

**Figure 1:**
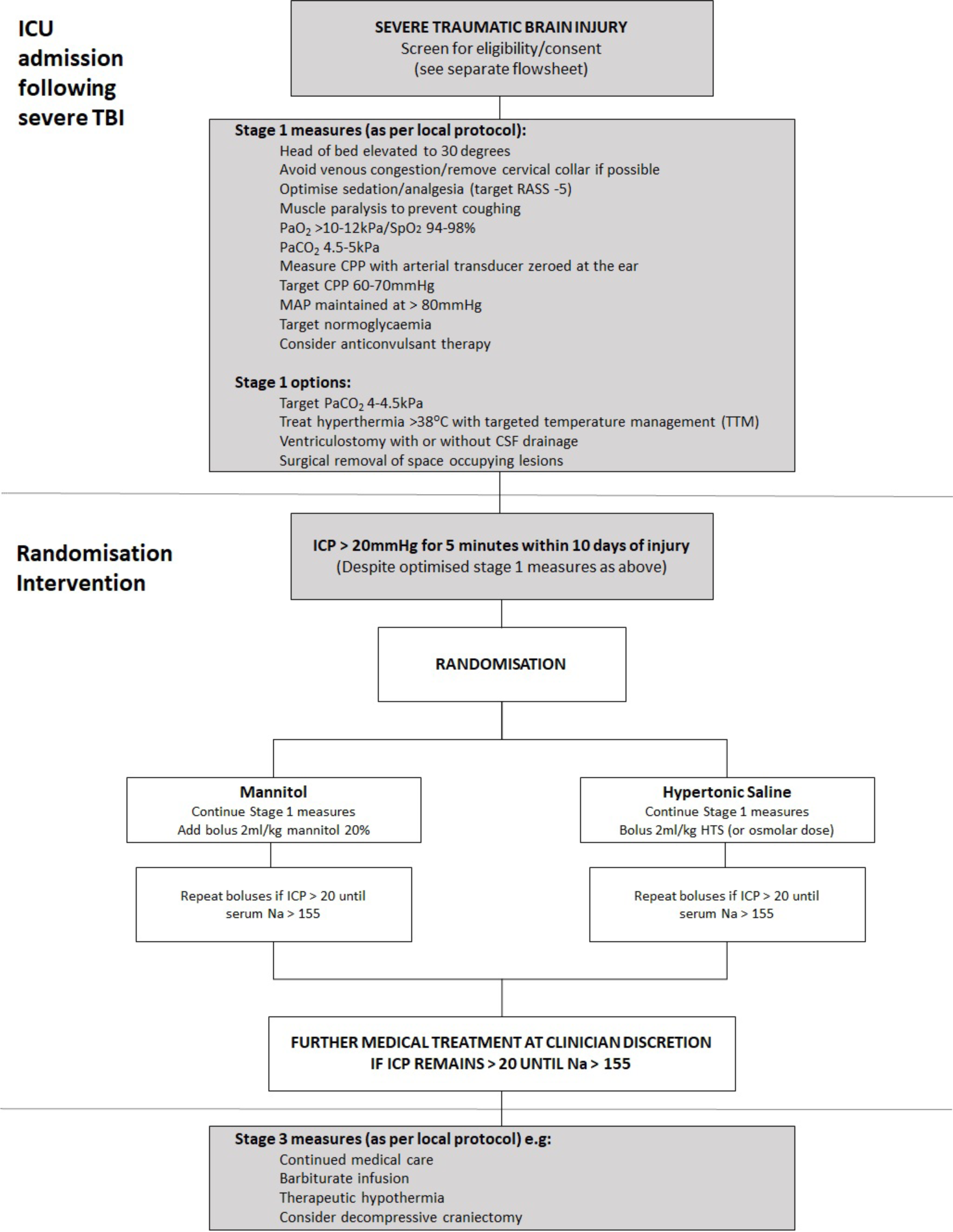
SOS Trial schematic.

Mortality will be reported from hospital records up until discharge and tracked after discharge using the NHS Digital tracking service, hospital records and GP records. ICU and hospital length of stay will be obtained from local centres.

### Statistical analysis

A detailed statistical analysis plan (SAP) will be developed during the trial, submitted to the DMEC for review and made publicly available prior to the database lock and commencement of analysis. The timing and frequency of the interim analyses will be discussed and agreed with the DMEC members and a detailed SAP will be written by the trial statistician and approved by the DMEC prior to any interim analysis. It is anticipated that no more than one formal interim analysis will take place during the course of the study. In making a decision to terminate the clinical trial, the DMEC will use the statistical evidence as guidance to their decision making and will be also presented with a 95% confidence interval of the treatment difference.

#### Primary outcome analysis

The primary analysis will be conducted on all outcome data obtained from all participants as randomised and regardless of protocol adherence, i.e. intention to treat analysis – unless a patient has specifically withdrawn their consent regarding the use of their data. A complier average causal effect (CACE) analysis will be used to address the issue of non-compliance.^19^ For the primary outcome analysis, the proportion of patients with favourable versus unfavourable 6-month GOS-Ewill be compared between the two intervention arms using the logistic regression model. This analysis will be adjusted for key clinically important co-variates. Odds ratios and 95% confidence intervals will be presented. As a supplementary analysis, the ordinal nature of the 8-point GOS-E questionnaire will also be assessed using appropriate ordinal regression models. Stacked bar charts will be used to display the ordinal GOS-E questionnaire data.

#### Secondary outcomes analyses

Survival status to hospital discharge will be examined in a similar way to the binary GOS-E questionnaire. In addition to this survival status over the course of the study (3, 6 and 12 months) and to time to discharge (ICU and hospital) will be assessed using Kaplan-Meier plots. The survival curves will be assessed using the log-rank test (unadjusted) and the Cox-proportional Hazards model (adjusted).

Continuous variables will be examined using linear regression models and summarised using mean, standard deviation, median and range values. Categorical data will be assessed using logistic regression models and summarised using the number of patients and proportions. Where appropriate, 95% confidence intervals will be presented with the appropriate point estimates.

#### Economic analyses

A detailed Health Economics Analysis plan (HEAP) will be developed during the trial, submitted to the DMEC for review and made publicly available prior to the database lock. Analysis will be conducted from an NHS and personal social services perspective, adhere to the recommendations of the NICE Reference Case.^20^ Resource use will include intervention, hospital (ICU, HDU and ward days) and community costs (primary care and social care costs) in the first 12 months following intervention. Resources will be costed using national reference unit costs where available, reflated to current prices. Health-related quality of life (EQ-5D-5L) responses will be used to generate quality-adjusted life years (QALYs) using the value set recommended by NICE at the time of analysis. Baseline EQ-5D-5L values will be imputed to reflect the unconscious health state and applied to all patients, minimising potential bias in the QALY AUC calculation.^21,22^

Within-trial analysis (to 12 months) using bivariate regression of costs and QALYs will inform a probabilistic assessment of the incremental cost-effectiveness ratio,^23^ cost-effectiveness acceptability and value-of-information of further research. Following best practice, missingness mechanisms will be explored and multiple imputation methods will be used within the analysis.^24^ Costs and outcomes arising during the trial will be undiscounted, reflecting the 12-month time horizon. Sensitivity and sub-group analyses will be pre-determined. More extensive economic modelling using decision-analytic methods may be considered, extending the time horizon and decision context, if costs and benefit profiles are non-convergent at 12 months.

#### Exploratory Bayesian analysis

In order to obtain more insight into the primary outcome, Bayesian methods will be used taking various information from the literature.

### Missing data

Every effort will be made to minimise missing baseline and outcome data in this trial. A further exploratory analysis to assess the impact of any missing outcome data on the GOS-E questionnaire will be examined using multiple imputation techniques.

### Data monitoring

The WCTU will be responsible for trial monitoring and visits will be conducted in accordance with the trial monitoring plan and will comply with the principles of Good Clinical Practice (GCP). On-site monitoring visits during the trial will check the accuracy of data entered into the clinical trial database against the source documents, adherence to the protocol, procedures and GCP, and the progress of patient recruitment and follow-up.

### Patient and public involvement

This application is informed through a series of meetings with survivors and carers of patients with previous brain injury, facilitated through Headway West Midlands (https://www.headway.org.uk). The group are supportive of this trial. Patient and public contributed to the trial proposal, protocol development and design of patient facing materials. We discussed and decided together our joint position on the use of placebo, trial outcomes and how to optimise the process for approaching, informing and consenting relatives (including the use of professional legal representative) and optimising follow-up. During the conduct of the trial two PPI members will join the trial management group and two will join the independent Trial Steering Committee.

We will follow INVOLVE best practice guidance in our approach. We will meet with the PPI group at the start of the study and regularly thereafter (monthly initially and then 3 monthly) to enable full involvement through the trial and have included funds to support this. We will work with our PPI group to ensure that we are all clear about expectations and jointly agree a role description, terms of reference and organisational responsibilities including payments. Our PPI leads will be readily accessible to the group. We will provide training and support through informal mentorship with experienced PPI and formal training through our CRN PPI group. The PPI group will help keep patients and public informed through the progress of the trial and lead the dissemination of the trial findings to lay persons.

### Safety, ethics and dissemination

#### Adverse outcomes

The occurrence of adverse events related to hyperosmolar therapy are poorly reported in the published clinical trials. The main anticipated potential adverse events are electrolyte disturbance and acute kidney injury. Adverse event/reactions will be assessed for seriousness and reported in accordance with MHRA guidelines.

#### Regulatory and ethics approvals

The East of England - Essex Research Ethics Committee (REC: 19/EE/0228 - flagged for trials involving clinical trials in patients without capacity) and Medical Healthcare Regulatory Authority (MHRA) (EudraCT:XXXXXXX) have approved the study protocol. The trial registration number is XXXXXXX. The study will comply with the principles for sharing clinical trial data from publicly funded clinical trials.

### Confidentiality

In order to maintain confidentiality, all CRFs, questionnaires, study reports and communication regarding the study will identify the patients by the assigned unique trial identifier and initials only. Patient confidentiality will be maintained at every stage and will not be made publicly available to the extent permitted by the applicable laws and regulations.

### Dissemination

The study will be reported in accordance with the Consolidated Standards of Reporting Trials (CONSORT) guidelines.^20^ The study findings will be presented at national and international meetings with abstracts on-line. Presentation at these meetings will ensure that results and any implications quickly reach all of the UK and international intensive care community. In accordance with the open access policies proposed by the NIHR, we aim to publish the clinical findings of the trial as well as a paper describing the cost-effectiveness in the NHS setting in high quality peer-reviewed open access (via Pubmed) journals. The NIHR HTA also requires that a detailed study report is published in the HTA journal. Finally, an on-going update of the trial will also be provided on the Warwick Clinical Trials Unit website and social media platforms e.g. Twitter (@sos_trial).

## Discussion

While mannitol has been the traditional hyperosmolar agent of choice in the management of raised ICP, use of HTS as first line therapy is increasing in the UK. The reasons for this shift remain unclear and may relate to perceived benefits in cardiovacscularly unstable polytrauma patients. However, what is clear is that there remains limited high quality trial evidence to support this change and potential for harm e.g. hypernatraemia. The SOS Trial has therefore been designed as an open and pragmatic clinical trial to determine the comparative clinical and cost effectiveness of mannitol vs HTS in adult patients with TBI and raised ICP. Results will also complement the French COBI trial comparing continuous hypertonic saline therapy against mannitol^21^ and will allow the possibility for an individual patient data network meta-analysis in the future.

The study protocol has been carefully designed with the support of clinicians in recruiting study centres to reflect and mirror ICP management pathways in routine clinical use. Through this, we hope to maximise patient recruitment and clinician engagement. As a range of concentrations of HTS are in use nationally to treat raised ICP, recruiting study centres will be allowed to use the standard concentration of HTS in use locally but in an equi-osmolar dose to match 2ml/kg of 3% HTS. We have also chosen an ICP threshold of 20mmHg for >5 minutes based on ICP management pathways acquired from recruiting study centres and previous large randomised controlled trials in this area.^14,22^ The sodium threshold for cessation of osmotherapy was set at 155mmol/l following discussions with experts in the field and recruiting centres. We acknowledge this is potentially lower than some clinicians would consider unsafe to proceed with further boluses of osmotherapy but feel this reflects routine clinical practice. However, we will monitor the effect this has on the study during the pilot stage of the study and review the threshold if required by the data monitoring committee or trial steering committee.

As with other trials in this clinical area, blinding to the intervention drugs was considered to be impractical and costly.^9^ This is because as highlighted, mannitol has a signature side effect causing a marked diuresis and hypertonic saline administration results in a rise in serum sodium levels. It would therefore not be possible to conceal treatment allocation without adversely affecting overall care of the patient (i.e. it would not be safe or practical to hide urine output or sodium levels in a critically ill patient with TBI) which is a potential limitation of the study design. To overcome this potential bias, the study design therefore includes blinded assessment of primary and other outcomes to minimise bias from knowledge of treatment assignment.

The inclusion of a placebo arm for the study was also carefully considered but also rejected for a number of reasons. In a survey of ICU, emergency medicine and neurosurgical clinicians conducted in advance of the HTA application, 80% of respondents were willing to randomise to mannitol or hypertonic saline but only 10% were willing to randomise to placebo. This would adversely affect enrolment and would risk contamination/cross over. This lack of equipoise is also consistent with the latest Brain Trauma Foundation guidelines which state “the committee is universal in its belief that hyperosmolar agents are useful in the care of patient with severe TBI”. It was also considered unlikely that the in the setting of current practice and beliefs, that the requirements of the EU Clinical Trials Directive could be fulfilled in demonstrating a “direct clinically relevant benefit” for placebo. Finally, discussions with patient and public partners involved in the trial, indicated there was discomfort regarding the inclusion of a placebo arm.

In conclusion, it remains unclear whether there is a treatment benefit to the use of either mannitol or hypertonic saline in the management of raised ICP after severe TBI. If results from the SOS Trial show the superiority of hypertonic saline over mannitol in improving neurological long term outcomes, then this has the potential to change clinical practice in the acute management of patients with raised ICP.

## Data Availability

This is a clinical trial protocol. All data is available in the manuscript.

## Contributions

GDP, MJR, TV, MW, CS, PD, DM, AK, PH, JM and RL conceived the study. All authors made a substantial contribution to the protocol development. RL is the study statistician. MJR, CS and GDP wrote the first draft of the manuscript and all authors critically revised the manuscript and gave final approval of the version to be published.

## Acknowledgements

The authors would like to acknowledge the support of the Warwick Clinical Trials Unit, the UK Critical Care Research Group, the Intensive Care Society, the NIHR National Critical Care Specialty Group, patient representatives and Headway, the Data Monitoring Committee and external members of the Trial Steering Group.

